# Hereditary hemorrhagic telangiectasia prevalence estimates calculated from gnomAD allele frequencies of predicted pathogenic variants in *ENG* and *ACVRL1*

**DOI:** 10.1101/2024.12.20.24319290

**Authors:** Anthony R. Anzell, Carter White, Brenda Diergaarde, Jenna C. Carlson, Beth L. Roman

## Abstract

**Background:** Hereditary hemorrhagic telangiectasia (HHT) is considered a fully penetrant autosomal dominant disorder characterized by the development of arteriovenous malformations. Up to 96% of HHT cases are caused by heterozygous loss-of-function mutations in *ACVRL1* or *ENG*, which encode proteins that function in bone morphogenetic protein signaling. HHT prevalence is estimated at 1 in 5000 and is accordingly classified as rare. However, HHT is suspected to be underdiagnosed due to variable age of onset and expressivity and lack of awareness of HHT among the medical community.

**Methods:** To estimate the true prevalence of HHT, we summed allele frequencies of predicted pathogenic variants in *ACVRL1* and *ENG* using three methods. For method one, we included Genome Aggregation Database (gnomAD v4.1) variants with ClinVar annotations of pathogenic or likely pathogenic, plus unannotated variants with a high probability of causing disease. For method two, we evaluated all *ACVRL1* and *ENG* gnomAD variants using threshold filters based on accessible *in silico* pathogenicity prediction algorithms. For method three, we developed a machine learning-based classification system to improve the classification of missense variants.

**Results:** Based on gnomAD variants, we calculated an HHT prevalence of between 2.1 in 5000 (method 1, most conservative) and 11.9 in 5000 (method 3, least conservative), or roughly 2 to 12-times higher than current estimates. Application of our machine learning-based classification method, which performed with over 97% accuracy, revealed missense variants as the greatest contributor to pathogenic allele frequency and similar HHT prevalence across genetic ancestries.

**Conclusions:** Our results support the notion that HHT is underdiagnosed and that HHT may not actually be a ”rare” disease.

## Introduction

Hereditary hemorrhagic telangiectasia (HHT) is an autosomal dominant vascular disorder characterized by the development of arteriovenous malformations (AVMs), which are direct connections between arteries and veins^1^. Small AVMs, or telangiectasias, typically form in the skin of the hands and face and the nasal and gastrointestinal mucosae, and rupture of mucosal telangiectases leads to recurrent hemorrhage and anemia. Larger AVMs may develop in the brain, lungs, and liver and can result in hemorrhagic or ischemic stroke, brain abscess, dyspnea, and high-output heart failure.

Heterozygous loss of function mutations in *ENG* (endoglin) and *ACVRL1* (encoding Activin-like kinase 1, ALK1) result in HHT1 and HHT2, respectively, and account for up to 96% of HHT cases^2–4^. *ENG* and *ACVRL1* encode proteins that function in bone morphogenetic protein (BMP) signaling in endothelial cells. Although the precise effect of pathway activation on endothelial cell gene expression and behavior is not fully understood, activity is clearly required for embryonic vascular development and AVM prevention^1^.

The reported prevalence of HHT in populations without founder effects has generally increased over time, likely because of heightened awareness and improved clinical diagnosis. For example, HHT prevalence was reported as 0.1 in 5000 in Northern England in 1990^5^ 0.3 in 5000 in Vermont, USA in 1990^6^; 0.8 in 5000 in County Fyn, Denmark in 1995^7^; and 1.6 in 5000 in Buenos Aires, Argentina in 2023^8^. Yet, based on the consensus of HHT medical experts, the true prevalence of HHT is thought to be considerably higher than suggested by clinically confirmed cases^9^. HHT clinical diagnostic criteria, referred to as the Curaçao criteria, include: 1) spontaneous and recurrent epistaxis, 2) multiple nasal, oral, and/or cutaneous telangiectasias, 3) visceral AVMs, particularly in the gastrointestinal tract, brain, lungs, and liver, and 4) family history^9, 10^. While presentation of at least three of these four straightforward criteria merits a definitive diagnosis of HHT, several factors contribute to HHT underdiagnosis. First, there is high variability in expressivity even within affected families^11^. Second, while penetrance is considered to be complete, it is age-related^5, 12^, likely due to the need for a somatic second hit in the wild type copy of the germline-mutated gene^13–15^. Accordingly, the Curaçao criteria are less effective in diagnosing children and young adults^16^. Third, the most common symptoms of HHT, nosebleeds and telangiectasias, are observed in the general population, albeit at lower frequency, and may not prompt physicians to suspect HHT^17^. Fourth, HHT and other rare diseases are not well emphasized in medical or dental curricula^18^, so provider awareness remains low.

The challenges in clinical diagnosis of HHT warrant an alternative approach to estimating prevalence. In HHT, disease-causing mutations have been observed across the entirety of *ACVRL1* and *ENG* and include missense, nonsense, and small and large indels affecting coding sequence, as well as splice site and intronic variants^19^. Given that HHT is dominant and considered to be fully penetrant, any variant that results in loss of protein function is expected to cause disease over the carrier’s lifespan. However, pathogenicity of many *ACVRL1* or *ENG* variants is difficult to establish; there are no standardized, low-cost functional assays of pathogenicity, resulting in designation of many as variants of unknown significance (VUS).

The public availability of large-scale genomic databases allows unbiased estimation of rare disease prevalence based on allele frequency of known or predicted disease-causing variants. ^20–22^ For example, ClinVar^23^ contains variants with accompanying pathogenicity classifiers based on clinical and/or functional analysis, whereas the Genome Aggregation Database (gnomAD v4.1)^24^ includes human variants derived from numerous whole-exome and whole-genome studies of mostly healthy populations. For gnomAD variants lacking ClinVar classification, one can query pathogenicity using, for example, the ENSEMBL Variant Effect Predictor (VEP)^25^, a user-friendly interface that aggregates publicly available prediction algorithms for variant classification.

In this work, we utilized data from gnomAD v4.1 to identify variants in *ACVRL1* and *ENG* and employed three different methods to calculate HHT prevalence. In method 1, we took a conservative approach and included only gnomAD variants that were ClinVar-annotated as pathogenic or likely pathogenic, plus frame shift, stop-gain, and splice site variants with a high probability of causing disease. In method 2, we employed *in silico* prediction algorithms available via gnomAD and Ensembl VEP to predict pathogenicity of all variants using simple threshold filters that require no coding skills. In method 3, we developed and optimized a robust ensemble random forest classification system that utilizes a combinatory approach of the best *in silico* prediction algorithms to improve pathogenicity prediction of missense variants and analyzed all other variant types by the threshold filter approach. Combining allele frequencies of predicted pathogenic variants for each of these methods, we calculate that HHT prevalence is somewhere between 2.1 in 5000 (method 1) and 11.9 in 5000 (method 3), or roughly 2-12 times higher than current global estimates of 1 in 5000^9^. Our data support the notion that HHT is underdiagnosed and suggest that HHT prevalence may be above the US Food and Drug Administration (FDA) threshold of 200,000 cases (or, < 3 in 5000) that currently defines a rare disease^26^.

## METHODS

Full methods are available in Supplemental Material. All data were extracted from publicly available databases, ClinVar and gnomAD v4.1. R functions written for evaluation of *in silico* pathogenicity prediction algorithm performance, machine learning model training, variant predictions, and prevalence and 95% confidence interval (CI) calculations are available in our GitHub repository (<u>PittRomanLab/Variant_Curation(github.com)</u>).

## RESULTS

### Method 1: HHT prevalence estimated by conservative curation of known and highly probable pathogenic variants

The prevalence of HHT is thought to be grossly underestimated due to missed diagnosis, but to our knowledge, no study has addressed the accuracy of this assertion from the perspective of genomic data analysis. To this end, we queried gnomAD v4.1 to identify *ENG* and *ACVRL1* variants. As a first-pass, and a conservative estimation of prevalence, we summed allele frequencies for gnomAD variants with a ClinVar annotation of likely pathogenic or pathogenic and at least a 1-star review rating (assertion criteria provided by at least one submitter), plus frameshift/stop-gain and splice site variants with a high probability of being deleterious (Figure 1A). For the latter, we applied Combined Annotation Dependent Depletion (CADD)^27^ and SpliceAI^28^ thresholds and then excluded variants with a homozygous allele count greater than 0 or allele frequency greater than 0.00004, consistent with the variant curation guidelines for HHT^29^.

**Figure 1.**
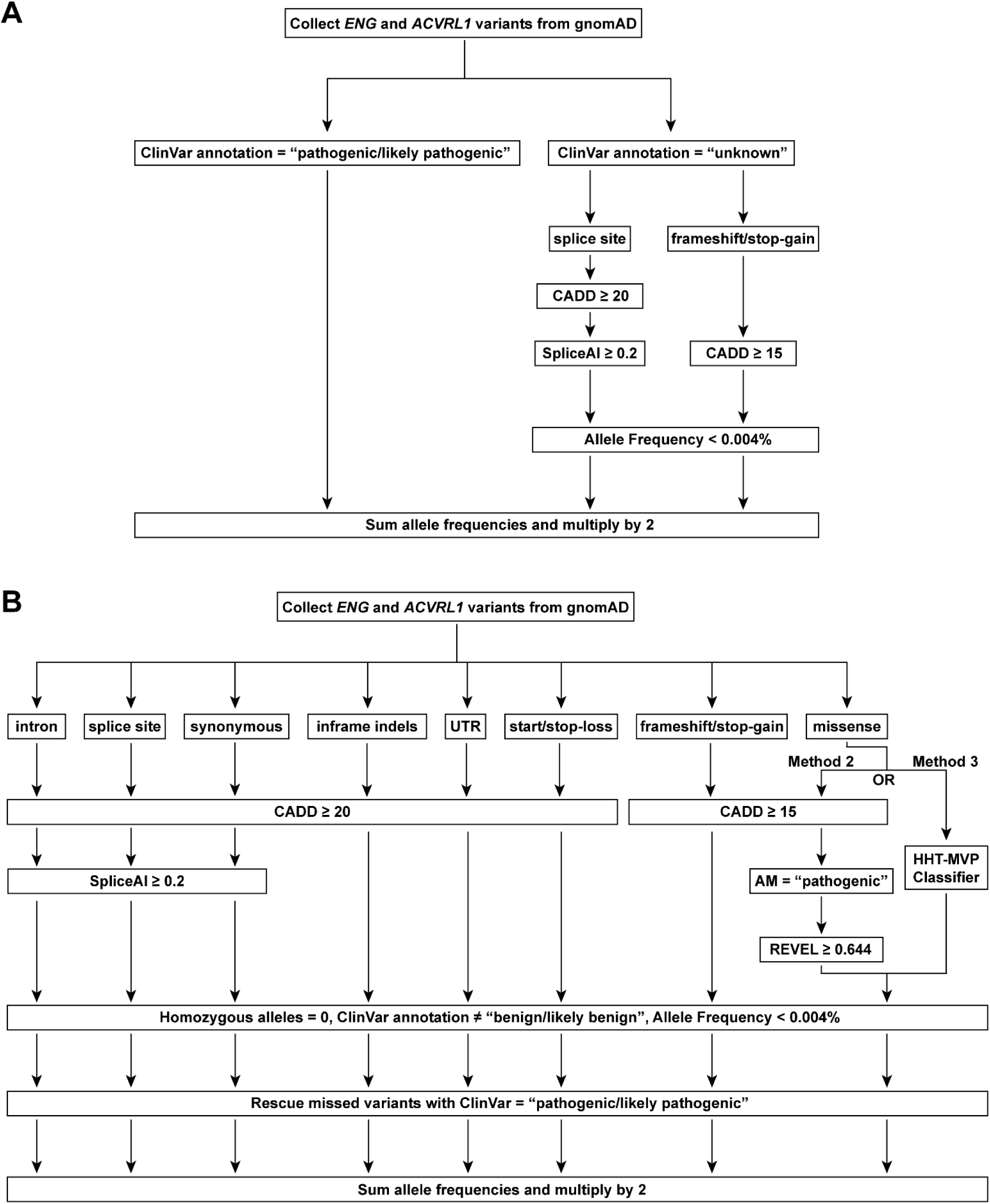
Flow charts for calculation of HHT prevalence from *ENG* and *ACVRL1* variants collected from gnomAD v4.1. A) Method 1: conservative curation. This method included only variants with a ClinVar annotation of pathogenic or likely pathogenic plus unannotated splice site and frameshift/stop-gain variants with a high probability of causing disease, as determined by SpliceAI, CADD, and allele frequency threshold filters. Allele frequencies were summed for known pathogenic and predicted pathogenic variants and the sum was multiplied by 2 to calculate disease prevalence per 5000. B) Methods 2 and 3: threshold filter-based and HHT-MVP curation. Variants were binned into 8 groups based on variant annotations. Missense variants were evaluated for pathogenicity using indicated threshold filters (method 2) or HHT-MVP (method 3). All other variants were evaluated for pathogenicity using the indicated threshold filters (method 2). Allele frequencies were summed for predicted pathogenic variants and the sum was multiplied by 2 to calculate disease prevalence per 5000.

For *ENG*, gnomAD 4.1 returned 2950 variants in total, the majority of which were intronic (1446; 48.9%) or missense (815; 27.6%) variants (Table 1 and Supplemental Excel File 1). Thirty-two of these variants had a ClinVar designation of likely pathogenic or pathogenic. We further retained 17 frameshift/stop-gain and 5 splice site variants that exceeded CADD and SpliceAI thresholds. Altogether, this method returned 54 variants: 2 intron, 12 splice site, 1 in-frame indel, 1 start loss, 31 frameshift/stop-gain, and 7 missense. Summing allele frequencies returned a pathogenic allele frequency for *ENG* of 0.0000753 (95% CI: 0.0000499, 0.000101), or an HHT1 prevalence of 0.8 in 5000 (95% CI: 0.5 in 5000 to 1.0 in 5000).

**Table 1:** HHT pathogenic allele frequencies and prevalence: Method 1, conservative curation.

| **ENG** |  |  |  |  |  |
| --- | --- | --- | --- | --- | --- |
| Variant Type | Total variants<br>in gnomAD<br>4.1 | ClinVar filter * | SpliceAI and/or<br>CADD threshold<br>filter † | Total variants | Allele frequencies |
| Intron | 1,446 | 2 | - | 2 | 2.052E-06 |
| Splice site | 127 | 7 | +5 | 12 | 2.303E-05 |
| Synonymous | 381 | 0 | - | 0 | 0 |
| In-frame indel | 13 | 1 | - | 1 | 6.841E-07 |
| 5' & 3' UTR | 132 | 0 | - | 0 | 0 |
| Start/Stop-Loss | 4 | 1 | - | 1 | 6.693E-07 |
| Frameshift or stop-gain | 32 | 14 | +17 | 31 | 3.590E-05 |
| Missense | 815 | 7 | - | 7 | 1.291E-05 |
| **ALL VARIANTS** | **2950** | **32** | **22** | **54** | **0.0000753**<br>(95% CI: 0.0000499,<br>0.000101) |
| **ACVRL1** |  |  |  |  |  |
| Variant Type | Total variants<br>in gnomAD<br>4.1 | ClinVar filter * | SpliceAI and/or<br>CADD threshold<br>filter † | Total variants | Allele frequencies |
| Intron | 747 | 0 | - | 0 | 0 |
| Splice site | 64 | 3 | +6 | 9 | 2.218E-05 |
| Synonymous | 309 | 0 | - | 0 | 0 |
| In-frame indel | 6 | 1 | - | 1 | 1.859E-06 |
| 5' & 3' UTR | 202 | 0 | - | 0 | 0 |
| Start/Stop Loss | 2 | 0 | - | 0 | 0 |
| Frameshift or stop-gain | 35 | 21 | +13 | 34 | 4.432E-05 |
| Missense | 484 | 21 | - | 21 | 6.241E-04 |
| **ALL VARIANTS** | **1849** | **46** | **19** | **65** | **0.000131**<br>(95% CI: 0.000106,<br>0.000156) |
| **Combined Totals (ENG + ACVRL1)** |  |  |  |  |  |
| **HHT allele frequency** |  |  |  | **0.000206** (95% CI: 0.000170, 0.000242) |  |
| **HHT prevalence** |  |  |  | **2.1 in 5000** (95% CI: 1.7 to 2.4 in 5000) |  |

\* ClinVar annotation of pathogenic or likely pathogenic with at least 1-start rating
† For splice site variants, CADD ≥ 20 and SpliceAI ≥ 0.2. For stop-gain/frameshift variants, CADD ≥ 15

For *ACVRL1*, gnomAD 4.1 returned 1849 variants in total, the majority of which were intronic (747; 40.4%) or missense (484; 26.2%) variants (Table 1). Forty-six of these variants had a ClinVar designation of likely pathogenic or pathogenic. We further retained 13 frameshift/stop-gain and 6 splice site variants that passed CADD and SpliceAI thresholds. Altogether, this method returned 65 variants: 9 splice site, 1 in-frame indel, 34 frameshift/stop-gain, and 21 missense. Summing allele frequencies returned a pathogenic allele frequency for *ACVRL1* of 0.000131 (95% CI: 0.000106, 0.000156), or an HHT2 prevalence of 1.3 in 5000 (95% CI: 1.1 in 5000 to 1.6 in 5000).

In summary, this conservative curation method returned a total HHT pathogenic allele frequency of 0.000206 (95% CI: 0.00170, 0.000242), corresponding to an HHT prevalence of 2.1 in 5000 (95% CI: 1.7 in 5000 to 2.4 in 5000), which is double the widely cited prevalence of 1 in 5000^9^ (Table 1). While most clinical studies find roughly equal prevalence of HHT1 and HHT2^4^, this method suggested that HHT2 is twice as common as HHT1. Accordingly, this method may undercount *ENG* pathogenic alleles.

### Method 2: HHT prevalence estimated by threshold filter-based curation of all gnomAD variants

To capture additional pathogenic variants, we next subjected all gnomAD *ENG* and *ACVRL1* variants to CADD, SpliceAI, Alpha Missense^30^, and Rare Exome Variant Ensemble Learner (REVEL)^31^ filters (Figure 1B and Supplemental Methods), using thresholds suggested for variant curation for HHT^29^. Data were further filtered by excluding variants with a homozygous allele count greater than 0, a ClinVar designation of benign or likely benign, allele frequency greater than 0.00004, or start-loss variants in *ACVRL1* (due to the presence of an in-frame downstream start codon), as per variant curation guidelines for HHT^29^ .

Application of CADD thresholds to the 2950 *ENG* variants retained 569 variants, the majority of which (510; 89.6%) were missense variants (Table 2 and Supplemental Excel File 1). For intronic, splice site, and synonymous variants, application of spliceAI retained a total of 17 variants. For missense variants, application of AlphaMissense followed by REVEL filters returned 12 variants. In total, the threshold filters returned 68 pathogenic variants, none of which were lost after application of the exclusion criteria. However, we failed to capture 8 variants with a ClinVar designation of pathogenic or likely pathogenic with assertion criteria provided, which we rescued in our final analysis. Ultimately, our final results included 7 intronic variants, 12 splice site variants, 3 in-frame indels; 5 UTR variants, 2 start-loss/stop-loss, 31 frameshift/stop-gain variants, and 16 missense variants. Summing allele frequencies returned a pathogenic allele frequency for *ENG* of 0.000163 (95% CI: 0.000125, 0.000202) or an HHT1 prevalence of 1.6 in 5000 (95% CI: 1.3 in 5000 to 2.0 in 5000).

**Table 2:** HHT pathogenic allele frequencies and prevalence: Method 2, threshold filter-based curation.

| **ENG** |  |  |  |  |  |  |  |  |  |
| --- | --- | --- | --- | --- | --- | --- | --- | --- | --- |
| Variant Type | Total variants in gnomAD 4.1 | Threshold filters |  |  |  | Excluded variants # | Missed variants** | Total variants | Allele frequencies |
|  |  | CADD | SpliceAI<sup>‡</sup> | AM<sup>§</sup> | REVEL |  |  |  |  |
| Intron | 1,446 | 7<sup>*</sup> | 6 | - | - | -0 | +1 | 7 | 6.972E-06 |
| Splice site | 127 | 13<sup>*</sup> | 11 | - | - | -0 | +1 | 12 | 2.303E-05 |
| Synonymous | 381 | 0<sup>*</sup> | 0 | - | - | -0 | +0 | 0 | 0 |
| In-frame indel | 13 | 2<sup>*</sup> | - | - | - | -0 | +1 | 3 | 8.046E-06 |
| 5' & 3' UTR | 132 | 5<sup>*</sup> | - | - | - | -0 | +0 | 5 | 3.027E-05 |
| Start/Stop-Loss | 4 | 2<sup>*</sup> | - | - | - | -0 | +0 | 2 | 1.392E-06 |
| Frameshift or stop-gain | 32 | 30<sup>†</sup> | - | - | - | -0 | +1 | 31 | 3.590E-05 |
| Missense | 815 | 510<sup>†</sup> | - | 78 | 12 | -0 | +4 | 16 | 5.781E-05 |
| **ALL VARIANTS** | **2950** |  | **(68)** |  |  | **(-0)** | **(+8)** | **76** | **0.000163**<br>(95% CI: 0.000125, 0.000202) |

| **ACVRL1** |  |  |  |  |  |  |  |  |  |
| --- | --- | --- | --- | --- | --- | --- | --- | --- | --- |
| Variant Type | Total variants in gnomAD 4.1 | Threshold filters |  |  |  | Excluded variants # | Missed variants** | Total variants | Allele frequencies |
|  |  | CADD | SpliceAI<sup>‡</sup> | AM<sup>§</sup> | REVEL |  |  |  |  |
| Intron | 747 | 6<sup>*</sup> | 3 | - | - | -0 | +0 | 3 | 2.056E-06 |
| Splice site | 64 | 9<sup>*</sup> | 9 | - | - | -0 | +0 | 9 | 2.218E-05 |
| Synonymous | 309 | 0<sup>*</sup> | 0 | - | - | -0 | +0 | 0 | 0 |
| In-frame indel | 6 | 2<sup>*</sup> | - | - | - | -0 | +0 | 2 | 5.300E-06 |
| 5' & 3' UTR | 202 | 0<sup>*</sup> | - | - | - | -0 | +0 | 0 | 0 |
| Start/Stop Loss | 2 | 1<sup>*</sup> | - | - | - | -1 | +0 | 0 | 0 |
| Frameshift or stop-gain | 35 | 34<sup>†</sup> | - | - | - | -0 | +0 | 34 | 4.432E-05 |
| Missense | 484 | 374<sup>†</sup> | - | 134 | 100 | -4 | +4 | 100 | 1.998E-04 |
| **ALL VARIANTS** | **1849** |  | **(149)** |  |  | **(-5)** | **(+4)** | **148** | **0.000274**<br>(95% CI: 0.000232, 0.000315) |

| **Combined Totals (ENG + ACVRL1)** |  |  |  |  |  |  |  |
| --- | --- | --- | --- | --- | --- | --- | --- |
| **HHT allele frequency** |  |  |  |  |  |  | **0.000437** (95% CI: 0.000380, 0.000494) |
| **HHT prevalence** |  |  |  |  |  |  | **4.4 in 5000** (95% CI: 3.8 to 4.9 in 5000) |

<sup>\*</sup>CADD ≥ 20 <sup>†</sup>CADD ≥ 15 <sup>‡</sup>SpliceAI ≥ 0.2 <sup>§</sup>AlphaMissense = likely pathogenic <sup>||</sup>REVEL ≥ 0.644 <sup>#</sup>Homozygous ≠ 0, ClinVar = benign or likely benign, allele frequency > 0.004%, or start-loss variant for ACVRL1 <sup>\*\*</sup>ClinVar = pathogenic or likely pathogenic with at least 1-star rating but not captured by threshold filters

Application of CADD thresholds to the 1849 *ACVRL1* variants retained 425 variants, the majority of which (374; 88%) were missense variants (Table 2 and Supplemental Excel File 1). For intronic, splice site, and synonymous variants, application of spliceAI retained a total of 12 variants. For missense variants, application of AlphaMissense followed by REVEL filters returned 100 variants. In total, the threshold filters returned 149 pathogenic variants, 5 of which were lost after application of exclusion criteria, leaving a total of 144 variants. Using this filtering method, we failed to capture 4 missense variants indicated as pathogenic or likely pathogenic with assertion criteria provided in ClinVar, which we rescued in our final analysis.

Ultimately, our final results included 3 intronic variants, 9 splice site variants, 2 in-frame indel; 34 frameshift/stop-gain variants, and 100 missense variants. Summing allele frequencies returned a pathogenic allele frequency for *ACVRL1* of 0.000274 (95% CI: 0.000232, 0.000315), or an HHT2 prevalence of 2.7 in 5000 (95% CI: 2.3 in 5000 to 3.2 in 5000).

In sum, this simple threshold filtering method returned a total HHT pathogenic allele frequency of 0.000437 (95% CI: 0.000380, 0.000494), corresponding to an HHT prevalence of 4.4 in 5000 (95% CI: 3.8 in 5000 to 4.9 in 5000), which is nearly 5 times the widely cited prevalence of 1 in 5000^9^ (Table 2). Similar to our conservative estimate from method 1, the pathogenic allele frequency for *ENG* is roughly half that of *ACVRL1* when using method 2.

### Evaluation of *in silic*o pathogenicity prediction algorithms for *ENG* and *ACVRL1*

#### missense variants

The threshold filtering scheme used in Method 2 relied on user-friendly *in silico* prediction algorithms to predict pathogenicity. However, these algorithms are designed for general use and are not gene specific. Therefore, we sought to evaluate the performance of multiple *insilico* prediction algorithms using all *ENG* and *ACVRL1* missense variants with a known ClinVar classification. The rationale for this endeavor was two-fold. First, we wanted to gauge accuracy of the accessible and user-friendly pathogenicity predictions programs that we used above (CADD, Revel, AlphaMissense), specifically with respect to HHT gene missense variants. Second, we wanted to identify additional algorithms that might have similar or higher predictive power to identify HHT pathogenic missense variants. To this end, we utilized ClinVar to generate a ground truth data set of annotated *ENG* and *ACVRL1* missense variants (Table S1, Supplementary Methods). There were 911 missense variants in ClinVar, 417 of which were annotated as benign or pathogenic. We further pruned this dataset to 386 variants (262 pathogenic, 124 benign) that had at least a 1-star rating (assertion criteria provided by at least one submitter) and produced an ENSEMBL VEP output. Utilizing the ground truth dataset, we analyzed each variant using 17 prediction algorithms to determine pathogenicity based on classification thresholds (see Supplementary Methods). For each algorithm, we compared predictions to the ClinVar classification, generated a confusion matrix (Figure S1A-R), and computed accuracy, sensitivity (ability to correctly classify known pathogenic variants), and specificity (ability to correctly classify known benign variants) (Figure 2A). In the combined dataset, the best performing algorithms were AlphaMissense, VEST4, and metaRNN, with accuracy > 90%, sensitivity > 87%, and specificity > 85%. By contrast, CADD performed with high sensitivity (98.4%) but low specificity (49.2%), whereas REVEL performed with lower sensitivity (83.9%) but high specificity (95.2%). These results are consistent with the fact that the CADD threshold filter retained more variants than AlphaMissense and REVEL filters (Table 2). The poorest performing algorithms, as assessed by accuracy, were PrimateAI (48.7%), MutationAssessor (56.2%), LRT (68.4%), and LIST.S2 (71.5%) (Figure 2A, Figure S1).

**Figure 2.**
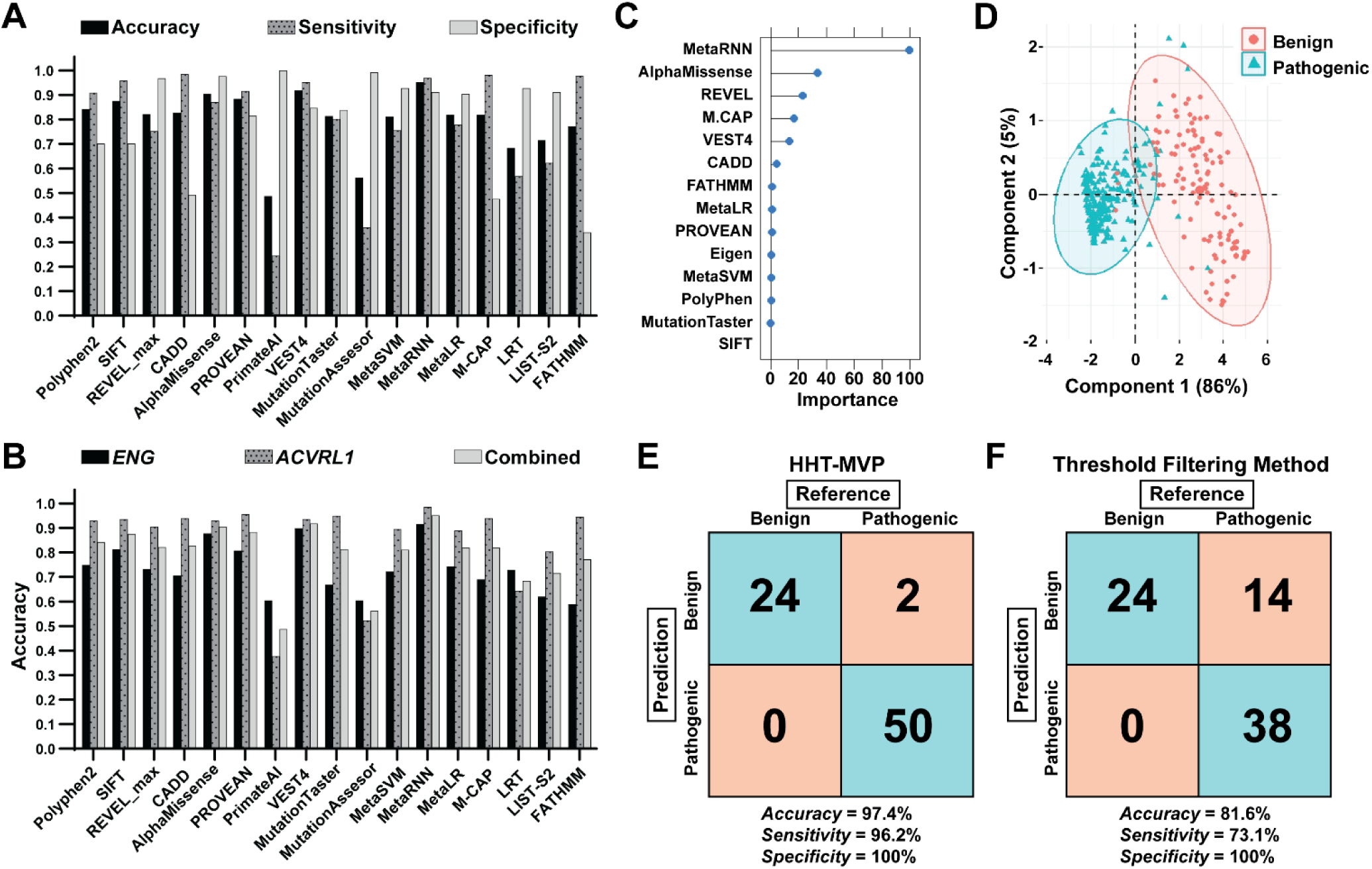
Evaluation of *in silico* pathogenicity prediction algorithm performance for HHT gene missense variants and machine learning development. A) Accuracy, sensitivity, and specificity of prediction algorithm performance using the ClinVar master dataset of known HHT pathogenic and benign missense variants in *ENG* and *ACVRL1*, combined. B) Accuracy of prediction algorithm performance of the ClinVar master dataset for missense variants, for *ENG, ACVRL1,* or combined. *ENG*: n = 187; 74 pathogenic, 113 benign. *ACVRL1*: n = 199; 188 pathogenic, 11 benign. Combined: n = 386; 262 pathogenic, 124 benign. C) Variable importance plot to determine the features used for prediction training. D) PCA of known ClinVar variants based on 6 most important features. E,F) Confusion matrix of missense variants within the test set predicted by HHT-MVP (E) or the threshold filtering method (F). n = 76; 52 pathogenic, 24 benign.

Generally, the prediction algorithms performed with higher accuracy for *ACVRL1* variants than *ENG* variants (Figure 2B). However, the accuracy for *ACVRL1* variants is likely artificially inflated due to the preponderance of pathogenic variants in the dataset (188 pathogenic, 11 benign). The accuracy of the prediction algorithms for classifying *ENG* variants (74 pathogenic, 113 benign) was similar to the combined group. AlphaMissense displayed a high accuracy of 87.7%, while REVEL had a relatively low accuracy of 73.3% and low sensitivity of 33.7% (Figure 2B, Figure S2). This likely accounts for the large dropout of *ENG* missense variants after applying the REVEL filter. These results highlight the need for careful analysis of *in silico* pathogenicity prediction algorithm performance for use in gene-specific variant classification.

Moreover, they suggest the need for a more sophisticated method for pathogenicity prediction than the threshold filtering method described above (Figure 1B, Table 2).

### Development of an ensemble machine learning-based pathogenicity classification system for HHT-related genes

#### Feature Selection

To develop a robust and accurate ensemble machine learning variant classification system for HHT genes, we selected the 13 highest performing prediction algorithms/features that returned over 80% accuracy when analyzing our ground truth data set of 386 missense variants (Figure 2): SIFT, PolyPhen, CADD, REVEL, AlphaMissense, PROVEAN, VEST4, MutationTaster, M.CAP, FATHMM, MetaRNN, MetaLR, and MetaSVM. In addition to these 13 features, we also used the Eigen prediction algorithm as a potential feature, which uses a supervised approach to derive the aggregate functional score from various annotation resources^32^. To train and test the machine-learning classification model, we used R coding language to randomly split our ground truth dataset into a training set (80%, 310 variants total; 210 pathogenic, 100 benign) and a test set (20%, 76 variants total; 52 pathogenic, 24 benign). Using the training set, we initially trained a random forest algorithm utilizing the R caret package^33^, which resulted in a high accuracy of 95.3% (Table S2). To prevent overfitting, we evaluated the importance of each feature using the varImp function in Caret to identify the features used by the random forest model to make predictions. The variable importance plot showed that MetaRNN, AlphaMissense, REVEL, M.CAP, VEST4, and CADD were the six main features used for prediction training (Figure 2C). To further visualize the separation of pathogenic and benign variants using these six features, we applied a principal component analysis (PCA) (Figure 2D). Pathogenic and benign variants aggregated in distinct populations with minimal overlap, demonstrating the ability of these features to accurately predict pathogenicity.

#### Evaluation of machine-learning algorithms

Ensemble machine learning consists of a “meta” approach that aims to increase model performance by combining the predictions of multiple models^34^. Although our random forest algorithm performed with an accuracy of 95.3%, we attempted to increase prediction performance using three additional types of machine learning algorithms: boosting, bagging, and stacking. We tested two boosting algorithms, C5.0 and Stochastic Gradient Boosting (GBM). Boosting uses ensemble algorithms to sequentially correct the predictions of prior models via generating a weighted average of prediction. We tested one additional bagging algorithm: Bagged Classification and Regression Trees (Bagged CART). Bagging averages predictions from multiple decision trees. Finally, we tested five stacking sub-models: Linear Discriminate Analysis (LDA), CART, Generalized Linear Model (GLM), k-nearest neighbor (kNN), and Support Vector Machine with a Radial Basis Kernel Function (svmRadial). Stacking generates a prediction based on the predictions of multiple types of sub-models^34^. Compared to the random forest model, all other algorithms performed with similar accuracy utilizing the training data (Table S2). The svmRadial sub-model marginally outperformed all other models with an accuracy of 96.6%. To determine if model performance could be further improved, we combined the individual stacking sub-models into a single meta-model using the “caretStack” function in the R Caret package^35^. Proper stacking of sub-models requires low correlation between the individual algorithms^34^, ensuring that each sub-model is uniquely skilled in its prediction performance. This allows the meta-model to make a prediction based on diverse informative decisions made by the sub-models. Because kNN had the lowest correlation with respect to the other algorithms (Table S3), we combined the predictions of each model from our training data using the kNN algorithm, which resulted in a 97.6% accuracy with a K = 5 (number of neighbors considered for classification) (Table S2). Lastly, we stacked the five sub-models using a random forest algorithm, combining all of the sub-model predictions to make a classification. This method, hereafter termed the HHT missense variant predictor (HHT-MVP), generated our most accurate model, with an accuracy of 97.7% (Table S2) and an mtry = 2 (corresponds to the number of input features a decision tree considers at any given point in time). Due to the high performance of HHT-MVP, no other parameters were tuned.

#### Verification of ensemble HHT-MVP model and comparison to filtering threshold method

We verified the accuracy of HHT-MVP utilizing the test dataset, which contained 76 variants (52 pathogenic, 24 benign) previously unseen by the algorithm. The accuracy of HHT-MVP was 97.4% (AUC = 0.981) (Figure 2E). Only two pathogenic variants were misclassified as benign, and no benign variants were misclassified as pathogenic, returning a sensitivity of 96.2% and a specificity of 100%. To compare performance between HHT-MVP and our threshold filtering method (method 2), we subjected our test dataset and the complete ground truth dataset to the three threshold filters used above: CADD ≥ 15, REVEL ≥ 0.644, and AlphaMissense = “likely pathogenic”. We compared the threshold filter output to the ClinVar reference prediction using a confusion matrix and calculated performance. The accuracy of the threshold filtering method with respect to the test dataset was 81.6% (∼16% reduction in accuracy) with 14 pathogenic variants being incorrectly predicted as benign variants (Figure 2F). No benign variants were misclassified as pathogenic, resulting in a sensitivity of 73.1% and a specificity of 100%. With respect to the larger ground truth dataset, the accuracy of the threshold filtering method was 85.2%, with 55 pathogenic variants being incorrectly predicted as benign variants and only 2 benign variants being classified as pathogenic, resulting in a sensitivity of 68.9% and a specificity of 99% (Figure S1S). These results demonstrate that HHT-MVP provides a greater accuracy in missense variant curation as compared to a standard threshold filtering approach. Additionally, it suggests that the threshold filtering method results in a conservative prevalence estimate: it efficiently filters out benign variants, but at the cost of filtering out a significant number of pathogenic variants. Accordingly, the true prevalence of HHT, based on gnomAD 4.1 variants, is likely underestimated by the threshold filtering method.

### Method 3: HHT prevalence estimated by HHT-MVP curation of missense variants and threshold filter-based curation of all other variants

HHT-MVP returned 153 and 204 pathogenic missense variants for *ENG* and *ACVRL1*, respectively. A total of 8 missense variants were lost after applying exclusion criteria, and just 2 ClinVar-annotated pathogenic missense variants were missed for *ENG* and 1 for *ACVRL1* (Table 3 and Supplemental Excel File 1). Compared to threshold filtering (method 2), HHT-MVP classified an additional 137 *ENG* and 104 *ACVRL1* missense variants as pathogenic (Table 2 and Table 3). Of missense variants with a ClinVar annotation of VUS, conflicting evidence, or no reference provided, 4-8% were classified as pathogenic by method 2, whereas ∼20-29% were classified as pathogenic by HHT-MVP (Table s). Summing pathogenic missense allele frequencies from HHT-MVP and pathogenic allele frequencies of all other variant types from method 2 returns a total pathogenic allele frequency for *ENG* of 0.000569 (95% CI: 0.000499, 0.000639), or an HHT1 prevalence of 5.7 in 5000 (95% CI: 5.0 in 5000 to 6.4 in 5000), and a pathogenic allele frequency for *ACVRL1* of 0.000622 (95% CI: 0.000566, 0.000679), or an HHT2 prevalence of 6.2 in 5000 (95% CI: 5.7 in 5000 to 6.8 in 5000). The similar frequencies are consistent with the observed similar prevalence of HHT1 and HHT2^4^. Summing *ENG* and *ACVRL1* pathogenic allele frequencies results in a total pathogenic allele frequency of 0.00119 (95% CI: 0.00110, 0.00128), suggesting an overall HHT prevalence of approximately 11.9 in 5000 (95% CI: 11.0 in 5000 to 12.8 in 5000). This prevalence is 2.69-times higher than predicted by the threshold filtering method and is nearly 12-times higher than current estimates based on clinical diagnoses.

**Table 3:** HHT pathogenic allele frequencies and prevalence: Method 3, HHT-MVP + threshold filter-based curation.

| **ENG** |  |  |  |  |  |  |  |
| --- | --- | --- | --- | --- | --- | --- | --- |
| Variant Type | Total variants in gnomAD 4.1 | HHT-MVP | Threshold Filter Method * | Excluded variants<sup>†</sup> | Missed variants<sup>‡</sup> | Total variants | Allele frequencies |
| Missense | 815 | 153 | - | -4 | +2 | 151 | 4.632E-04 |
| All other variants | 2135 | - | 56 | -0 | +4 | 60 | 1.056E-04 |
| **ALL VARIANTS** | **2950** |  |  |  |  | **211** | **0.000569**<br>(95% CI: 0.000499, 0.000639) |
| **ACVRL1** |  |  |  |  |  |  |  |
| Variant Type | Total variants in gnomAD 4.1 | HHT-MVP | Threshold Filter Method * | Excluded variants<sup>†</sup> | Missed variants<sup>‡</sup> | Total variants | Allele frequencies |
| Missense | 484 | 204 | - | -4 | +1 | 201 | 5.485E-04 |
| All other variants | 1364 | - | 49 | -1 | +0 | 48 | 7.386E-05 |
| **ALL VARIANTS** | **1849** |  |  |  |  | **249** | **0.000622**<br>(95% CI: 0.000566, 0.000679) |
| **Combined Totals (ENG + ACVRL1)** |  |  |  |  |  |  |  |
| **HHT allele frequency** |  |  |  |  |  | **0.00119** (95% CI: 0.00110, 0.00128) |  |
| **HHT prevalence** |  |  |  |  |  | **11.9 in 5000** (95% CI: 11.0 to 12.8 in 5000) |  |

<sup>\*</sup> Pathogenic variants as per threshold filter method, Table 2
<sup>†</sup> Homozygous ≠ 0, ClinVar = benign or likely benign, allele frequency > 0.004%, or start-loss variant for *ACVRL1*
<sup>‡</sup> ClinVar = pathogenic or likely pathogenic with at least 1-star rating but not captured by applied method

### HHT gene predicted pathogenic allele frequencies are similar across all genetic ancestries

Although HHT was originally thought to be most prevalent in European-descent populations^36^, this notion likely stemmed from ascertainment bias. To assess the distribution of HHT, we utilized the genetic ancestry data provided in gnomAD v4.1 to evaluate the allele frequency of all HHT-MVP-predicted pathogenic variants in multiple populations: African/African American, Admixed American (North and South Americas), Ashkenazi Jewish, East Asian, South Asian, European (including Finnish), European (excluding Finnish), Middle Eastern, and Remaining

(all other samples not categorized with the other groups). Of 807,162 sequences in gnomAD v4.1, the majority (662,057; 82.0%) are European due to inclusion of genetic data from the UK biobank ^37^. Most other ancestries are similarly represented with ∼15,000 - 50,000 sequences, although Middle Eastern ancestry is underrepresented with 3,031 (Table 4). Despite the variance in sample counts, the combined allele frequencies of all predicted pathogenic variants are similar across all genetic ancestries (0.001 - 0.0018, Table 4). Pathogenic *ACVRL1* and *ENG* allele frequencies are similar to each other in Admixed American and European populations, whereas, based on limited data, one or the other dominates in other populations (Table 4). These results are consistent with the notion that the HHT prevalence is similar regardless of genetic ancestry.

**Table 4:**
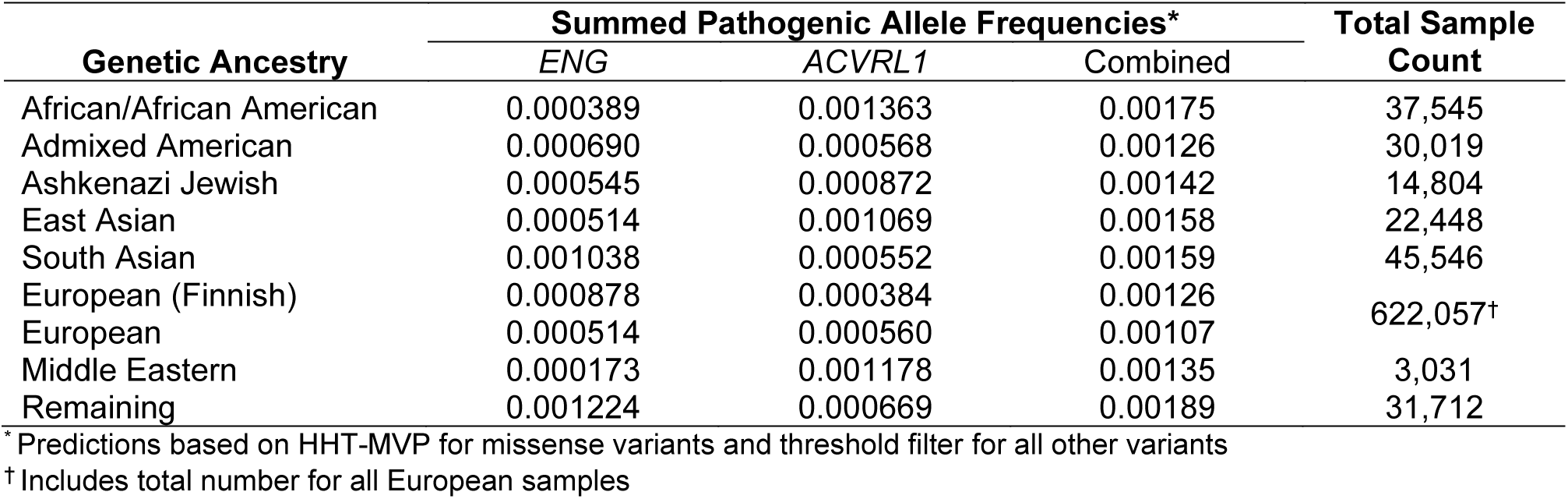
*ENG* and *ACVRL1* pathogenic allele frequencies across genetic ancestries.

## Discussion

The current estimate of HHT prevalence, based on clinical diagnosis, is 1 in 5000 people^9^. However, the disease is believed to be highly underdiagnosed due to variability in expressivity and age of onset coupled with insufficient awareness of this disease among clinical providers^5^. In this work, we leveraged gnomAD to identify variants in the HHT-related genes, *ACVRL1* and *ENG*, predict pathogenicity, and calculate the at-risk population for developing HHT. Using a conservative approach that captured only known and highly probable pathogenic variants and two less conservative methods—a threshold approach and a machine learning-based classification approach—we estimate that HHT prevalence is between 2.1 to 11.9 in 5000.

Accordingly, HHT may be 2-to-12 times more prevalent than the current estimates and may not be a “rare” disease.

In this work, we focused on variants in *ENG* and *ACVRL1*, which account for up to 96% of HHT cases^4^ . Disease-causing mutations are overwhelmingly private (that is, unique to a given family) and have been documented across the entirety of both of these genes^19^. According to ClinVar, *ENG* frameshift and nonsense mutations leading to premature termination codons (PTC) are most common in HHT1, whereas *ACVRL1* missense and PTC-causing mutations show similar frequency in HHT2 (Figure 3A). While conservative curation (method 1) returned similar results—an unsurprising outcome given that this was largely based on ClinVar annotations—the threshold filtering without or with HHT-MVP missense classification (methods 2 and 3, respectively) returned progressively increasing proportions of missense variants and decreasing proportions of PTC-causing variants (Figure 3A). Venn diagrams of all pathogenic variants or missense pathogenic variants collected by our three methods are concentric circles (Figure 3B). That is, all variants in the conservative curation set are included in the threshold filtering set, and all variants in the threshold filtering set are included in the threshold filtering plus HHT-MVP set. The threshold method captured 105 more variants than the conservative method, 88 of which were missense variants. The HHT-MVP captured 245 more missense variants than the threshold method. In ClinVar, roughly 57% and 40% of *ENG* and *ACVRL1* missense variants, respectively, are annotated as VUS. Given the higher accuracy of the HHT-MVP (97.4%) versus the threshold method (81.6%) with respect to missense variant classification, these data suggest that 1) the HHT-MVP classification system could improve classification of current missense VUS in *ENG* and *ACVRL1*; 2) pathogenic missense variants in both genes may be more prevalent than previously thought, and 3) both proteins may be sensitive to folding defects that lead to endoplasmic reticulum retention and associated degradation, as previously suggested^39–41^.

**Figure 3.**
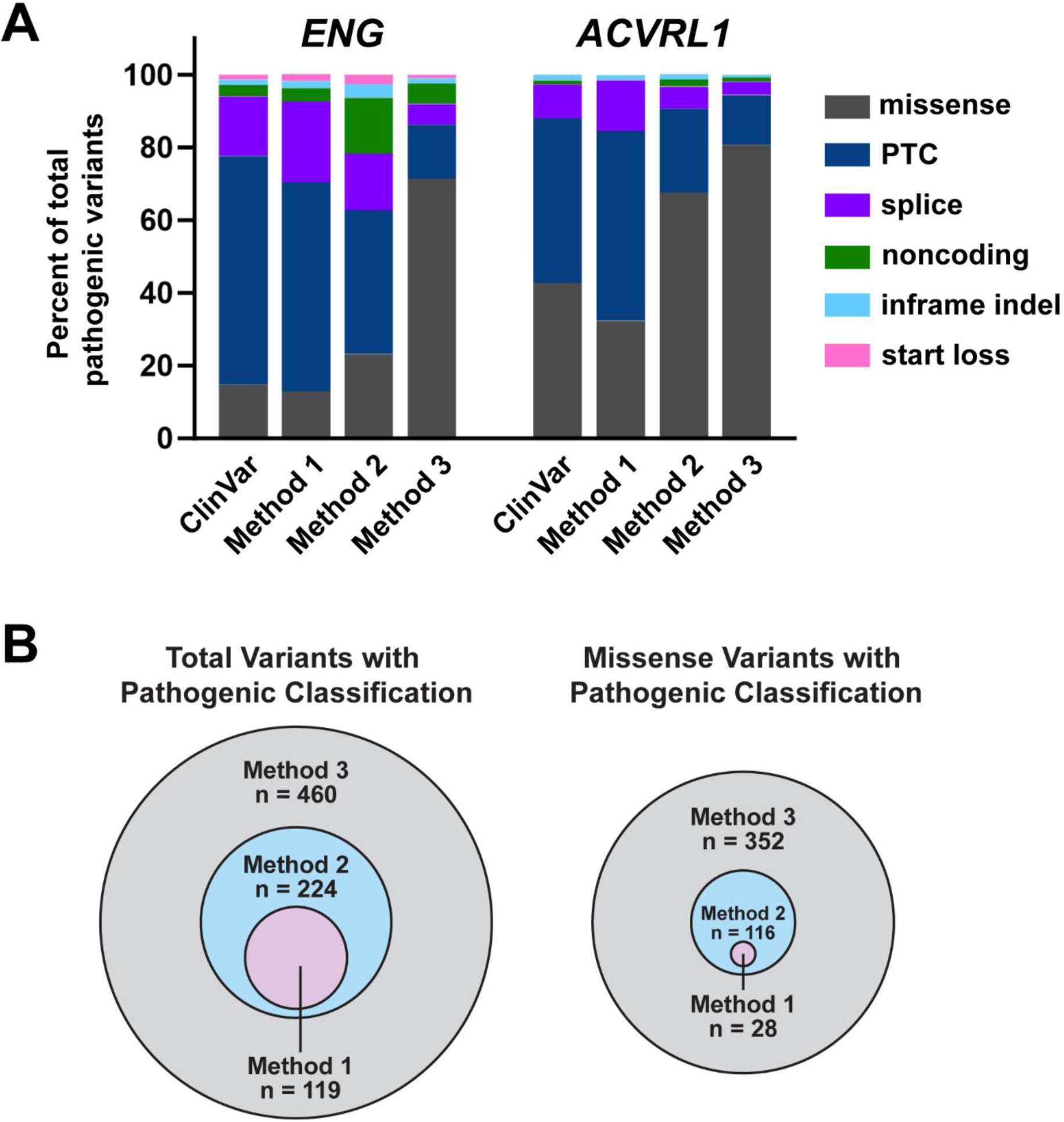
Comparison of pathogenic variants returned from three pathogenicity prediction methods. A) Pathogenic *ENG* and *ACVRL1* variants, grouped by variant type, as recorded in ClinVar and as designated by our three pathogenicity prediction methods. Data are presented as percent of total pathogenic variants. PTC = premature termination codon, including nonsense and frameshift variants. B) Venn diagrams of total pathogenic variants (left) and missense pathogenic variants (right) returned from the three pathogenicity prediction methods. Method 1: conservative curation. Method 2: threshold filtering curation. Method 3: threshold filtering + HHT-MVP curation.

Outside of *ENG* and *ACVRL1*, a very small percentage of HHT cases is caused by mutations in *SMAD4* ^4, 42^. However, of 298 pathogenic or likely pathogenic variants (excluding copy number variants) in ClinVar, just 64 are linked to HHT, with the remainder associated with other conditions, without HHT. Accordingly, using *SMAD4* genomic data to estimate HHT prevalence is unsupported. There are a few reports of mutations in *GDF2*, which encodes the ALK1 ligand, BMP9, causing HHT or an HHT-like syndrome^43, 44^; however, there is just one example of a clinically confirmed HHT family with a *GDF2* mutation^45^. In sum, although *SMAD4* and *GDF2* variants were excluded from our analysis, we would not expect a significant increase in HHT prevalence with the inclusion of these genes.

For autosomal dominant diseases, prevalence calculations based entirely on genomic data assume complete penetrance. HHT satisfies this criterion, with age-related penetrance of at least one manifestation of HHT approaching 100% by 40 years of age^5, 11, 12^. Although some studies suggest haploinsufficiency as a disease mechanism for HHT^46, 47^, others provide evidence that lesions are seeded by a somatic mutation in the wild type copy of the germline-mutated gene^13–15^. Age-related penetrance supports the latter mechanism, with genetic insults accumulating with age. Animal models of HHT also support this mechanism: AVMs develop with complete penetrance only in the homozygous and not the heterozygous state^48, 49^, and *Acvrl1* or *Eng* null endothelial cells can seed mosaic lesions in a wild type background^50, 51^.

Accordingly, we expect that germline heterozygosity for any pathogenic variant will result in some disease phenotype in nearly all carriers by age 40. Leveraging databases like All of Us^52^ or the UK Biobank^53^, in which electronic health records are coupled to genomic data, could add credence to our prediction method. Based on presumed underdiagnosis of HHT, we would expect that patients with pathogenic variants in HHT genes may be more likely to be diagnosed with common HHT symptoms (for example, epistaxis, anemia, GI bleed) than with HHT.

Despite the increasing volume of publicly available whole exome and whole genome sequencing, classification of rare genomic variants remains a major challenge to genetic diagnosis of rare Mendelian diseases. *In silico* prediction algorithms, which consider factors such as inter-species conservation and predicted or proven effects on protein structure and/or function^54^, provide insight into pathogenicity of genomic variants. However, choosing which prediction algorithm(s) to apply for any given gene is not clear-cut. For example, our threshold method resulted in a significant dropout of *ENG* missense variants after consecutive application of CADD, AlphaMissense, and REVEL filters, reducing likely pathogenic variants from 469 to 75 to 12, respectively. Yet, performance assessment revealed that AlphaMissense displayed a much higher accuracy than REVEL with respect to both HHT genes, suggesting the use of 3D structure predictions may be a more appropriate computational tool for HHT genes in variant curation. Regardless, it is clear that application of a threshold method is prone to error and likely underestimates prevalence, as evidenced by the low accuracy (85.2%) and sensitivity (68.9%) of this method when applied to a ClinVar ground truth dataset of *ENG* and *ACVRL1* missense variants of known pathogenicity.

To increase accuracy of pathogenicity prediction of missense variants, we tested the individual performance of 17 prediction algorithms on the ClinVar master data set and used the 13 best performing features to develop and train a machine learning algorithm, from which we identified six main features driving prediction. Using these features, we developed an ensemble machine learning-based classification model that derives power from the iterative process of evaluating each variant and weighing the predictions of multiple *in silico* algorithms to make the most accurate classification. However, as with all machine learning algorithms, there are inherent limitations stemming from training data. Our ClinVar ground truth dataset is balanced with respect to gene (187 *ENG,* 199 *ACVRL1*); however, it is unbalanced with respect to variant classification, with roughly 32% benign and 68% pathogenic variants. This skewing is driven primarily by *ACVRL1*, with 94% pathogenic variants and 6% benign variants. To ensure our model performance was not compromised by the imbalanced ground truth dataset, we employed the synthetic minority over-sampling technique (SMOTE) to synthetically balance the ground truth dataset^55^. We trained the same type of machine-learning algorithm using the SMOTE master dataset, which produced similar numbers with respect to accuracy of the test set and number of classified pathogenic missense variants (data not shown). Access to a larger training dataset of confirmed pathogenic and benign variants of all types is critical to enhancing the performance of HHT-MVP and extending its scope to other variant types.

Despite limitations, our HHT-MVP classification system performed with an accuracy of 97.4%, a sensitivity of 96.2%, and a specificity of 100% on a test set of 76 HHT gene variants (ClinVar designations: 52 pathogenic, 24 benign), representing a major improvement over the threshold filtering method due largely to the improved retention of pathogenic variants and reclassification of VUS (Figure 3 and Table S4). Accordingly, the HHT prevalence estimate, based on gnomAD variants, is roughly 2.7 times higher with the HHT-MVP classification system (11.9 in 5000) than with the threshold method (4.4 in 5000).

Out of the 915 *ENG* and *ACVRL1* missense variants in ClinVar^23^, about half (406) are VUS. In the absence of pedigree data, converting these VUS to pathogenic or benign designations requires functional validation. Conceptually, functional testing using patients’ own induced-pluripotent stem cell (iPSC)-derived endothelial cells provides an ideal approach^56^. However, this method is high cost, labor intensive, and low throughput. Moreover, given the likely requirement for a somatic second hit for lesion development^13^, decreased signaling is likely to be undetectable in the heterozygous state^57^. In vitro assays do exist for testing *ACVRL1* and *ENG* variant function^39, 58^, but they rely on transfection of immortalized non-endothelial cells, are low throughput, and are impractical for clinical testing. In this regard, HHT-MVP may serve as a useful tool to identify likely pathogenic variants that should proceed to functional validation.

Based exclusively on *in silico* analysis, we estimate that the prevalence of HHT, irrespective of genetic ancestry, is between 2.1 to 11.9 in 5000 people, which equates to ∼3.4 to 19.4 million people worldwide. This HHT prevalence estimate is at minimum 2 times to potentially 12 times the current estimate of 1 in 5,000 people^9^, supporting the notion that HHT is underrecognized and underdiagnosed and suggesting that HHT may not be a rare disease. In 2024, the National Institutes of Health (NIH) provided ∼$5.4 million in funding for 15 projects focused on HHT. By comparison, the NIH provided $35.4 million for 65 projects focused on the rare bleeding disorder, hemophilia (1 in 5,000 males)^59^. Accordingly, from a public health perspective, increased investment in HHT, which causes considerable medical and psychosocial morbidity^60, 61^, would be of enormous benefit.

## Data Availability

All data were extracted from public databases and are included in a supplemental excel file. ClinVar and gnomAD v4.1. R functions written for evaluation of in silico pathogenicity prediction algorithm performance, machine learning model training, variant predictions, and prevalence and 95% confidence interval (CI) calculations are available in our GitHub repository.

https://github.com/PittRomanLab/Variant_Curation

## Abbreviations

[*ACVRL1* (gene)]: Activin A receptor like type 1
[ALK1 (protein)]: Activin receptor-like kinase 1
(AM): AlphaMissense
(AVMs): arteriovenous malformations
(Bagged CART): Bagged Classification and Regression Trees
(BMP): bone morphogenetic protein
(CADD): Combined Annotation Dependent Depletion
(CSV): comma separated value
(FP): False Positive
(FN): False Negative
(GLM): Generalized Linear Model
(gnomAD): Genome Aggregation Database
(HHT): Hereditary Hemorrhagic Telangiectasia
(HHT-MVP): HHT missense variant predictor
(HGVS): Human Genome Variation Society
(indel): insertion/deletion
(kNN): k-nearest neighbor
(LDA): Linear Discriminate Analysis
(PAH): Pulmonary Arterial Hypertension
(REVEL): Rare Exome Variant Ensemble Learner
(gbm): Stochastic Gradient Boosting
(svmRadial): Support Vector Machine with a Radial Basis Kernel Function
(TP): True Positive
(TN): True Negative
(UTR): untranslated region
(VEP): Variant Effect Predictor
VUS: (variant of unknown significance)

## Acknowledgements

We would like to thank R. Minster and D. Weeks (University of Pittsburgh, PA, USA) for guidance on study design and project execution.

