## Supplemental Material for "Hereditary hemorrhagic telangiectasia prevalence estimates calculated from gnomAD allele frequencies of predicted pathogenic variants in *ENG* and *ACVRL1*"

### METHODS

#### Variant collection from gnomAD

The Genome Aggregation Database (gnomAD v4.1) was used to collect variants in the HHT genes, *ACVRL1* and *ENG.* Version 4.1 contains 730,947 exomes and 76,215 genomes (GRCh38). Each gene was searched on the gnomAD website (https://gnomad.broadinstitute.org) and variants were exported to a comma separated value (CSV) file. Variants that did not pass gnomAD's quality control process were not included in our study. Variant data were further processed using Microsoft Excel (Version 2406, Build 17726.20126, Redmond WA) or Rstudio software (Version 2024.04.0 Build 735, Boston MA). Rstudio software was used for all R computation.

#### ClinVar ground truth dataset for missense variants

For evaluation of in silico pathogenic prediction algorithms and machine learning model development, we used ClinVar (https://www.ncbi.nlm.nih.gov/clinvar/, downloaded May 2024) to collect known pathogenic and benign variants in *ACVRL1* and *ENG*. For each gene, we selected missense variants categorized as "Pathogenic", "Likely pathogenic", "Benign", and "Likely Benign" with at least a 1-star rating (assertion criteria provided by at least one submitter) for a total 398 variants. The number of *ACVRL1* and *ENG* variants for each category is listed in Table S1. Variants were exported to a single CSV file and used as our "ground truth dataset" for evaluation of variant pathogenicity prediction.

#### Feature extraction

Missense variants collected from gnomAD and ClinVar were run through the Ensembl Variant Effect Predictor (VEP) web interface[25] (Version 112, GRCh38.p14). Variants were inputted using Human Genome Variation Society (HGVS) nomenclature. Transcript database was set to Ensembl/GENCODE transcripts. Identifiers included gene symbol and transcript version. For the variant and frequency data section, "find co-located known variants" was set to no. For additional annotations, MANE was selected, "upstream/downstream distance (bp)" was set to 5,000, and IntAct was set to disabled. Under predictions, SIFT and PolyPhen were set to "Prediction and score", AlphaMissense was checked, SpliceAI was disabled, and dbNSF was enabled and the listed fields were chosen: LRT_score, LRT_converted_rankscore, LRT_pred, LRT_Omega, MutationTaster_score, MutationTaster_converted_rankscore, MutationTaster_pred, MutationTaster_model, MutationTaster_AAE, MutationAssessor_score, MutationAssessor_rankscore, MutationAssessor_pred, FATHMM_score, FATHMM_converted_rankscore, FATHMM_pred, PROVEAN_score, PROVEAN_converted_rankscore, PROVEAN_pred, VEST4_score, VEST4_rankscore, MetaSVM_score, MetaSVM_rankscore, MetaSVM_pred, MetaLR_score, MetaLR_rankscore, MetaLR_pred, Reliability_index, MetaRNN_score, MetaRNN_rankscore, MetaRNN_pred, M-CAP_score, M-CAP_rankscore, M-CAP_pred, REVEL_score, REVEL_rankscore, MutPred_score, MutPred_rankscore, MutPred_protID, MutPred_AAchange, MutPred_Top5features, MVP_score, MVP_rankscore, gMVP_score, gMVP_rankscore, MPC_score, MPC_rankscore, PrimateAI_score, PrimateAI_rankscore, PrimateAI_pred, DEOGEN2_score, DEOGEN2_rankscore, DEOGEN2_pred, BayesDel_addAF_score, BayesDel_addAF_rankscore, BayesDel_addAF_pred, BayesDel_noAF_score, BayesDel_noAF_rankscore, BayesDel_noAF_pred, ClinPred_score, ClinPred_rankscore, ClinPred_pred, LIST-S2_score, LIST-S2_rankscore, LIST-S2_pred, VARITY_R_score, VARITY_R_rankscore, VARITY_ER_score, VARITY_ER_rankscore, VARITY_R_LOO_score, VARITY_R_LOO_rankscore, VARITY_ER_LOO_score, VARITY_ER_LOO_rankscore, Aloft_Fraction_transcripts_affected, Aloft_prob_Tolerant, Aloft_prob_Recessive, Aloft_prob_Dominant, Aloft_pred, Aloft_Confidence, CADD_raw, CADD_raw_rankscore, CADD_phred, CADD_raw_hg19, CADD_raw_rankscore_hg19, CADD_phred_hg19, DANN_score, DANN_rankscore, fathmm-MKL_coding_score, fathmm-MKL_coding_rankscore, fathmm-MKL_coding_pred, fathmm-MKL_coding_group, fathmm-XF_coding_score, fathmm-XF_coding_rankscore, fathmm-XF_coding_pred, Eigen-raw_coding, Eigen-raw_coding_rankscore, Eigen-phred_coding, Eigen-PC-raw_coding, Eigen-PC-raw_coding_rankscore,

Eigen-PC-phred_coding. No further filtering options were chosen and "show all results" was selected for restrict results section.

VEP predictions were exported into Excel and variants were filtered by transcript ID (*ACVRL1*: ENST00000388922.9 and *ENG*: ENST00000373203.9) to return one prediction score per variant submitted. For the ground truth dataset, the VEP output was pasted into the ground truth dataset and variants were aligned by the "Upload_variation" number (VEP output) and the HGVS ID (ClinVar output) in Excel. For gnomAD dataset, after machine learning classification, the VEP output was joined to gnomAD data through HGVS ID alignment using RStudio. VEP output was further cleaned to remove unnecessary columns. Retained features included: SIFT, PolyPhen, am_class, am_pathogenecity, CADD_phred, Eigen.raw_coding, FATHMM_converted_rankscore, LIST.S2_score, LRT_pred, LRT_converted_rankscore, LRT_score, REVEL_rankscore, REVEL_score, VEST4_rankscore, VEST4_score, PrimateAI_pred, PrimateAI_score, PrimateAI_rankscore, PROVEAN_converted_rankscore, PROVEAN_score, MutationTaster_pred, MutationTaster_score, MutationTaster_converted_rankscore, MutationAssessor_pred, MutationAssessor_rankscore, MutationAssessor_score, MetaSVM_pred, MetaSVM_rankscore, MetaSVM_score, MetaRNN_pred, MetaRNN_rankscore, MetaRNN_score, MetaLR_pred, MetaLR_rankscore, MetaLR_score, M.CAP_score, M.CAP_rankscore, and M.CAP_pred.

**Evaluation of in silico pathogenicity prediction algorithms**

To evaluate the performance of PolyPhen, SIFT, REVEL_max, CADD, AlphaMissense (AM), PROVEAN, PrimateAI, VEST4, MutationTaster, MutationAssessor, MeatSVM, metaRNN, MetaLR, M.CAP, LRT, LIST2, and FATHMM in correctly identifying pathogenic variants, we employed our ClinVar ground truth dataset and calculated confusion matrices on predicted outcomes versus known outcome for each algorithm. Pathogenicity was determined for each variant using the following thresholds: REVEL_max > 0.644, SIFT < 0.05, PolyPhen >= 0.446, CADD_phred >= 15, VEST4_rankscore > 0.5, FATHMM_converted_rankscore >= 0.453, MutationAssessor_rankscore > 0.8, PrimateAI_pred = "D", am_class = "likely_pathogenic",PROVEAN_converted_rankscore >= 0.54382, MutationTaster_converted_rankscore >=0.52043, MetaRNN_rankscore >= 0.6149, MetaLR_pred = "D", MetaSVM = "D", M.CAP_pred = "D", LRT_pred = "D", LIST.S2_score >= 0.85. All variants predicted as pathogenic were assigned a "1", while benign variants were assigned a "0". Confusion matrixes were calculated using the R Caret package[33] and accuracy, sensitivity, and specificity were reported. The threshold cutoff for REVEL_max was set based on variant curation guidelines for HHT[29]. CADD score cutoff was determined arbitrarily by evaluating scores of known pathogenic and benign missense variants collected in ClinVar. All other threshold scores were determined by the dbNSFP database (dbNSFP README file, version 4.5c)[62-64].

**Machine Learning Classification**

Machine learning-based classification of missense variants was performed in R computing language using the Caret package[33]. Utilizing the ground truth dataset obtained from ClinVar that had been annotated using Ensembl VEP, we split the data randomly into a training set (80%) and a test set (20%) using the createDataPartition function. Several algorithms were individually tested for prediction accuracy including Boosting (C5.0 and Stochastic Gradient Boosting), Bagging (Random Forest and Bagged CART), and Stacking (Linear Discriminant Analysis (LDA), Classification and Regression Trees (CART), Logistic Regression (GLM), k-Nearest Neighbors (kNN), and Support Vector Machine with a Radial Basis Kernel Function (SVM)) models. Each of these algorithms were trained with the training set with 30 repetitions and performance was assessed via accuracy and kappa. The final model used for classification was composed of a bade model, consisting of the 5 stacking algorithms listed above, and a random forest meta model (final model termed "HHT-missense variant predictor (HHT-MVP)"). The test set was run through the final model to assess accuracy of an unknown dataset. Confusion matrices were produced to assess the performance of the model.

For classification of missense variants collected from gnomAD with ensemble VEP data, the data were imported into Rstudio and run through the trained final model using the predict function. Pathogenic or benign predictions were then exported with the original gnomAD/VEP data into a CSV file and compiled in Microsoft Excel with the other variant data.

**Collection of ClinVar variants for proportion comparisons across methods**

To compare variant proportions across methods and to pathogenic/likely pathogenic annotated ClinVar variants, we exported all pathogenic or likely pathogenic *ENG* and *ACVRL1* variants in the ClinVar database that were assigned a 1-star rating (assertion criteria provided by at least 1 submitter). Variants were then manually annotated with a molecular consequence based on RefSeq NM annotation and protein change information. Data were compiled in Excel and percentages of each variant type were compared to predicted pathogenic gnomAD variants.

**Collection of NIH Reporter stats**

For comparison of research dollars and number of funded projects between HHT and Hemophilia, NIH reporter was used. “Hereditary Hemorrhagic Telangiectasia” or “Hemophilia” was entered into the search bar. Data were then filtered by fiscal year 2024. Data were exported, including abstract, to excel. To ensure NIH projects were specific for HHT and Hemophilia, data were filtered using a “contains” filter in the abstract column for either “Hereditary Hemorrhagic Telangiectasia”/”HHT” or “Hemophilia”. The “total cost” column was summed using the subtotal function in Excel to calculate the total NIH research dollars awarded.

**Statistical Analysis**

Statistical analysis was performed in R computing language using Rstudio. Confusion matrices were produced to calculate accuracy, sensitivity, and specificity. Accuracy was defined as the ratio of total correct instances to the total instances:

$$Accuracy = \frac{True\ Positive\ (TP) + True\ Negative\ (TN)}{TP + TN + False\ Positive\ (FP) + False\ Negative\ (FN)}$$

Sensitivity or the True Positive Rate was defined as the measure of the model’s ability to correctly identify positive instances (pathogenic variants):

$$Sensitivity = \frac{TP}{TP + FN}$$

Specificity or the True Negative Rate was defined as the measure of the model’s ability to correctly identify negative instances (benign variants):

$$Specificity = \frac{TN}{TN + FP}$$

Graphs were generated using GraphPad Prism (Version 10.2.3) and R computing language.

**Prevalence and 95% confidence interval calculations**

In gnomAD 4.1, allele frequency is calculated for each variant by dividing its allele count by the total allele number. This denominator may vary over the length of a given gene due to differences in sequencing coverage at any given genomic position[65]. Note that allele frequency does not represent the percentage of individuals that carry a particular allele because each individual carries 2 alleles at all loci.

Because HHT is autosomal dominant with assumed complete penetrance, we calculated prevalence using the following equation:

$$y = \hat{p}(2)(5000)$$

Where $y =$ the number affected individuals in 5000, $\hat{p} =$ the summed allele frequencies of $m$ predicted pathogenic variants (i.e., $\hat{p} = \sum_{i=1}^{m} \hat{p}_i$) and 2 = the number of alleles per person. To calculate 95% CI surrounding $\hat{p}$ (summed pathogenic allele frequencies), the following formulas were used to calculate the lower and upper bounds:

$$lcb = \hat{p} - z * s$$

and

$$ucb = \hat{p} + z * s$$

Where $lcb$ = lower confidence bound, $ucb$ = upper confidence bound, $s$ = the standard error, and $z$ = the critical value. The standard error was calculated using the following formula:

$$s = \sqrt{\sum_{i=1}^{m} \frac{\hat{p}_i(1 - \hat{p}_i)}{n_i}}$$

Where $\hat{p}_i$ = allele frequency of variant $i$ (i.e., $\frac{allele\ count}{allele\ number}$) and $n_i$ = allele number of variant $i$. The critical value, $z$, is the $\left(1 - \frac{\alpha}{2}\right)^{th}$ quantile from a standard normal distribution.

### SUPPLEMENTAL TABLES

#### Table S1: Variants collected and filtered for ground truth dataset.

| Filtering Steps | Combined | *ENG* | *ACVRL1* |
|---|---|---|---|
| Total missense variants in ClinVar | 911 | 518 | 393 |
| Selected “Pathogenic”, “Likely Pathogenic”, “Benign”, and “Likely Benign” | 417 | 197 | 220 |
| At least 1 star | 398 | 193 | 205 |
| Produced VEP outputs | 386 | 187 | 199 |
| Pathogenic/Likely Pathogenic | 262 | 74 | 188 |
| Benign/Likely Benign | 124 | 113 | 11 |

#### Table S2: Performance of ensemble machine learning algorithms.

| Model | Accuracy | Kappa† |
|---|---|---|
| **Bagging** | | |
| Random Forest* | 0.953 | 0.892 |
| Random Forest | 0.956 | 0.898 |
| Bagged CART | 0.955 | 0.896 |
| **Boosting** | | |
| C5.0 | 0.957 | 0.899 |
| GBM | 0.958 | 0.903 |
| **Stacking sub-models** | | |
| svmRadial | 0.966 | 0.923 |
| LDA | 0.958 | 0.902 |
| CART | 0.957 | 0.898 |
| GLM | 0.952 | 0.888 |
| kNN | 0.941 | 0.863 |
| **Meta Ensemble** | | |
| kNN stack | 0.976 | 0.943 |
| HHT-MVP | 0.977 | 0.946 |

*Initial random forest algorithm trained using 14 features. All other algorithms were trained using 6 main features.

†Kappa is the comparison between observed and expected accuracy.

Bagged CART = Bagged Classification and Regression Trees, GBM = Stochastic Gradient Boosting, svmRadial = Support Vector Machine with a Radial Basis Kernel Function, LDA = Linear Discriminate Analysis, GLM = Generalized Linear Model, kNN = kNearest Neighbor.

**Table S3: Correlation analysis of stacking sub models.**

| | LDA | CART | GLM | kNN | svmRadial |
|---|---|---|---|---|---|
| **LDA** | 1 | 0.721299 | 0.950748 | 0.63039 | 0.746803 |
| **CART** | 0.721299 | 1 | 0.7299 | 0.537689 | 0.557713 |
| **GLM** | 0.950748 | 0.7299 | 1 | 0.692321 | 0.681713 |
| **kNN** | 0.63039 | 0.537689 | 0.692321 | 1 | 0.458972 |
| **svmRadial** | 0.746803 | 0.557713 | 0.681713 | 0.458972 | 1 |

LDA = linear discriminate analysis, CART = Classification and Regression Trees, GLM = Generalized Linear Model, kNN = k-Nearest Neighbor, svmRadial = Support Vector Machine with a Radial Basis Kernel Function

**Table S4: Capture rates of *ENG* and *ACVRL1* missense variants grouped by their ClinVar annotation in gnomAD v4.1.**

| ClinVar Annotation | Total Variants* | Pathogenic by Threshold Filter (n, (%))† | Pathogenic by HHT-MVP (n, (%))† |
|---|---|---|---|
| Pathogenic/ Likely Pathogenic | 29‡ | 20 (68.9%) | 26 (89.7%) |
| VUS | 186 | 7 (3.8%) | 41 (22.0%) |
| Conflicting Evidence | 54 | 4 (7.4%) | 11 (20.4%) |
| Not Available | 946 | 77 (8.1%) | 271 (28.6%) |

* Does not include missense variants with a CinVar designation of benign or likely benign.
† Variants that did meet inclusion criteria are not included in these data.
‡ Total number of variants with ClinVar annotation of pathogenic or likely pathogenic and a 1-star rating in ClinVar.

### SUPPLEMENTAL FIGURES

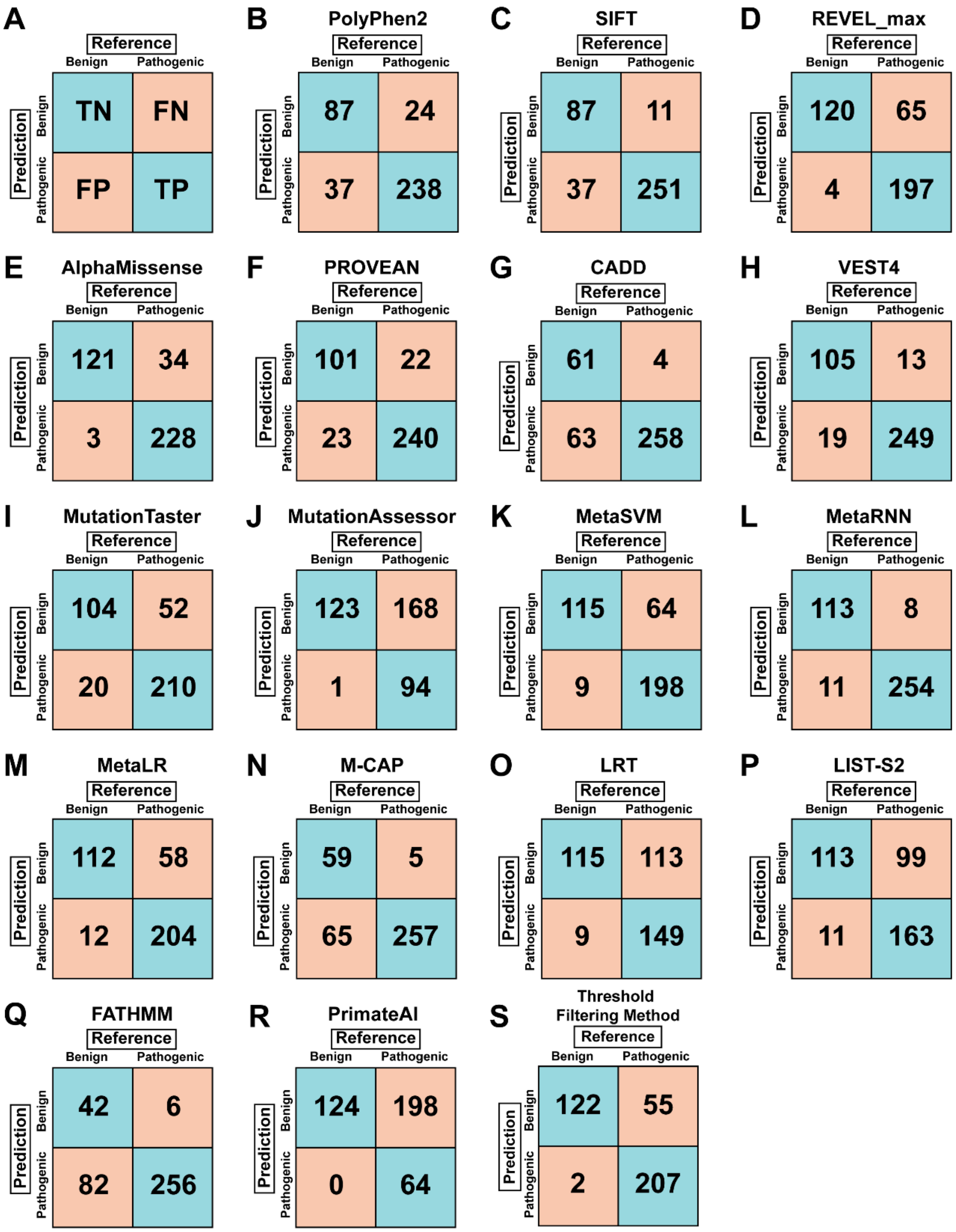


**Figure S1. Evaluation of in silico pathogenicity prediction algorithms for *ACVRL1* and *ENG* missense variants.** (**A**) Example confusion matrix indicating true negative (TN), false negative (FN), true positive (TP), false positive (FP) based on comparison of ClinVar (reference) and algorithm-predicted designation of benign or pathogenic. **(B-R)** Confusion matrices for 17 individual *in silico* pathogenicity prediction algorithms tested on a ground truth data set consisting of 262 pathogenic and 124 benign *ENG* and *ACVRL1* missense variants. **(S)** Confusion matrix for the threshold filtering method tested on a ground truth data set consisting of 262 pathogenic and 124 benign *ENG* and *ACVRL1*missense variants.

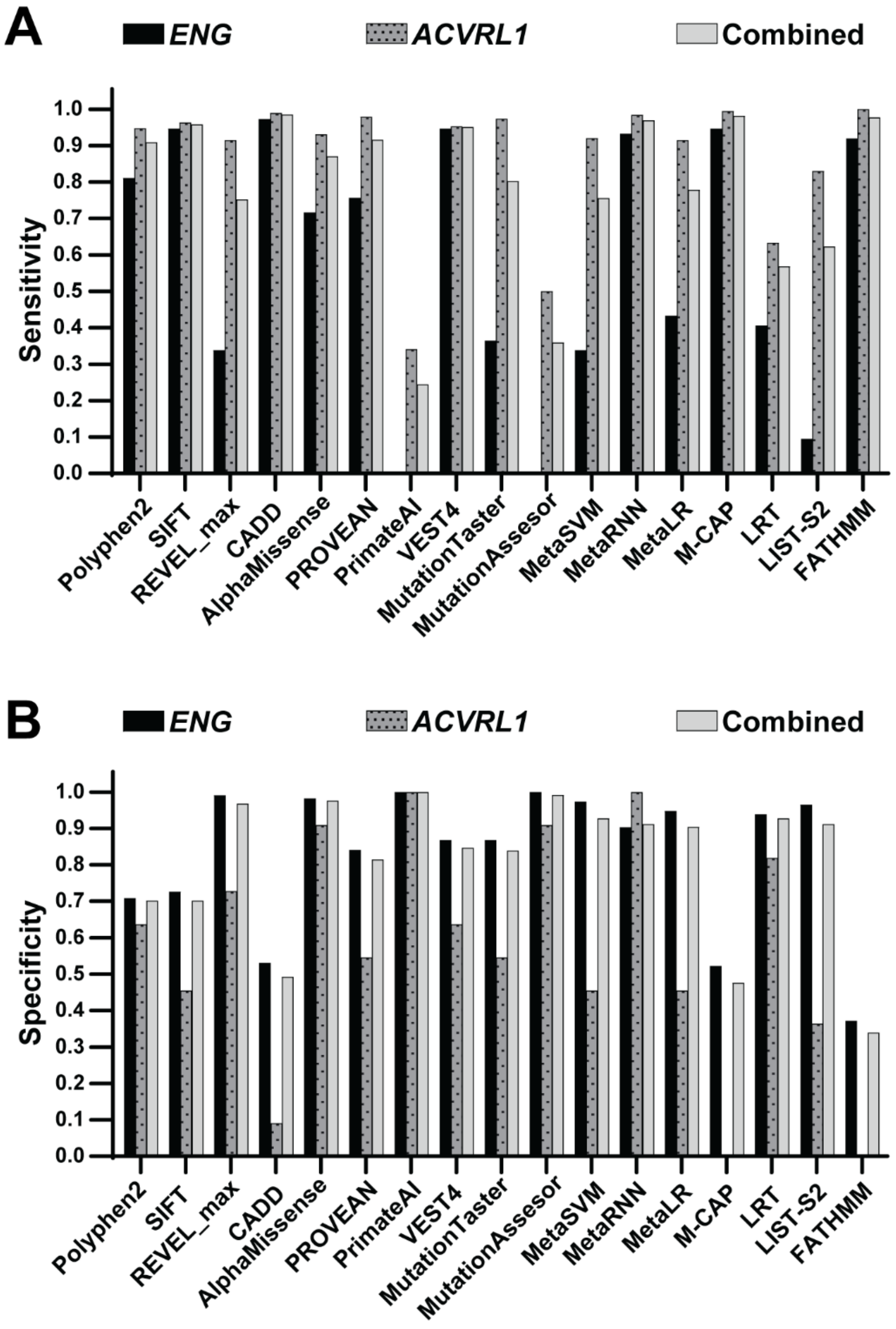


**Figure S2. Prediction performance of *in silico* pathogenicity algorithms.** Sensitivity (**A**) and specificity (**B**) of algorithm performance of the ClinVar ground truth dataset for each gene (*ENG* and *ACVRL1*). *ENG*: n = 187 (74 pathogenic, 113 benign); *ACVRL1*: n = 199 (188 pathogenic, 11 benign); combined: n = 386 (262 pathogenic, 124 benign).
